# Classification of COVID-19 in intensive care patients: towards rational and effective triage

**DOI:** 10.1101/2020.04.09.20058909

**Authors:** Xiaofan Lu, Yang Wang, Taige Chen, Yongsheng Li, Jun Wang, Fangrong Yan

**Affiliations:** State Key Laboratory of Natural Medicines, Research Center of Biostatistics and Computational Pharmacy, China Pharmaceutical University, Nanjing, China; Department of Radiology, The Affiliated Nanjing Drum Tower Hospital of Nanjing University Medical School, Nanjing, China; Medical School of Nanjing University, Nanjing, China; Department of Intensive Care Medicine, Tongji Hospital, Tongji Medical College, Huazhong University of Science and Technology, Wuhan, China; Department of Intensive Care Medicine, The First Affiliated Hospital of Soochow University, Suzhou, China

## Abstract

The number of pertinent researches of COVID-19 has increased rapidly but they mainly focused on the description of general information of patients with confirmed infection. We aimed to bridge the gap between disease classification and clinical outcome in intensive care patients, data of which are scarce and such classification could help in individual evaluation and provide effective triage for treatment and management. Specifically, we collected and filtered out 151 intensive care patients with complete medical records from Tongji hospital in Wuhan, China. We constructed a fully Bayesian latent variable model for integrative clustering of six data categories, including demographic information, symptoms, original comorbidities, vital signs, blood routine tests and inflammatory marker measurements. We identified four prognostic types of COVID-19 in intensive care patients, presenting a stepwise distribution in age, respiratory condition and inflammatory markers, suggesting the prognostic efficacy of these indicators. This report, to our knowledge, is the first attempt of dealing with classification of COVID-19 in intensive care patients. We acknowledge the limitation of ignoring the effect of treatment, but we believe such classification is enlightening for better triage, allowing for a more rational allocation of scarce medical resources in a resource constrained environment.

## Introduction

Severe acute respiratory syndrome coronavirus 2 (SARS-CoV-2) pneumonia is a newly recognized illness that has spread rapidly around the world. Previous studies on coronavirus disease 2019 (COVID-19) mainly described the general epidemiological, clinical, and radiological features of patients with confirmed infection (1). Little attention has been paid to the clinical characteristics and outcomes of intensive care patients with COVID-19, data of which are scarce but are of paramount importance to reduce mortality. We aimed to bridge the gap between disease classification and clinical outcome in intensive care patients, which could help in individual evaluation and provide effective triage for treatment and management.

## Methods

Data of 306 intensive care patients were obtained from Tongji hospital in Wuhan, China. Patients were hospitalized and admitted to intensive care wards from January 25 to February 25, 2020, who had been diagnosed with infection of SARS-CoV-2 according to WHO interim guidance. Data on the day of admission were collected, including six data categories: demographic information of age and gender, symptoms ([>10%] fever, fatigue, dry cough, anorexia, myalgia, dyspnea, expectoration, diarrhea), original comorbidities ([>5%] hypertension, diabetes, cardiovascular disease [CVD], chronic obstructive pulmonary disease [COPD], malignancy), vital signs (respiratory rate, heart rate, blood pressure, SpO_2_, FiO_2_), blood routine tests (count of white blood cell [WBC], lymphocyte, neutrophil, platelet and monocyte, red cell distribution width [RDW]) and inflammatory marker measurements (high-sensitivity C-reactive protein [hs-CRP], interleukin-2 receptor [IL-2R], IL-6, IL-8, IL-10, tumor necrosis factor-α [TNF-α]). Clinical outcome was 28-day mortality after admission to intensive care wards. The Ethics Commission of Tongji hospital approved this study, with a waiver of informed consent for the rapid emergence of this epidemic.

A total of 151 intensive care patients with complete medical records were further filtered out for this study. We constructed a fully Bayesian latent variable model for integrative clustering of the six data categories with different distributions (2). Specifically, age was dichotomized with 65-year cutoff according to the literature (3). Outliers were detected by *Tukey’s* method which uses interquartile (IQR) range approach, and outliers ranged above and below the 1.5×IQR were substituted with third quartile+1.5×IQR and first quartile-1.5×IQR, respectively. Data magnitude was manually checked and logarithmic transformation was utilized as appropriate; continuous data were further median-centered and scaled for being comparable. Binomial and Gaussian distributions were considered for categorical and continuous variables, respectively, and prior probability for the indicator variable gamma was set to 0.5 for each data category. Appropriate number of clusters was determined by minimizing the Bayesian information criterion. Only features with high posterior probability (*e*.*g*., 0.8) were kept. All statistical analyses were conducted with R3.6.2 using a Fisher’s exact test for categorical data and a Kruskal–Wallis test for multiple group comparison; Survival rates was generated by Kaplan-Meier curve and analyzed with log-rank test. For unadjusted comparisons, a two-sided *P*<0.05 was considered statistically significant.

## Results

We identified four types of intensive patients with COVID-19, which was tightly associated with clinical outcome (**Figure**). Characteristics of the four types were described below (**Table**).

**Figure.**
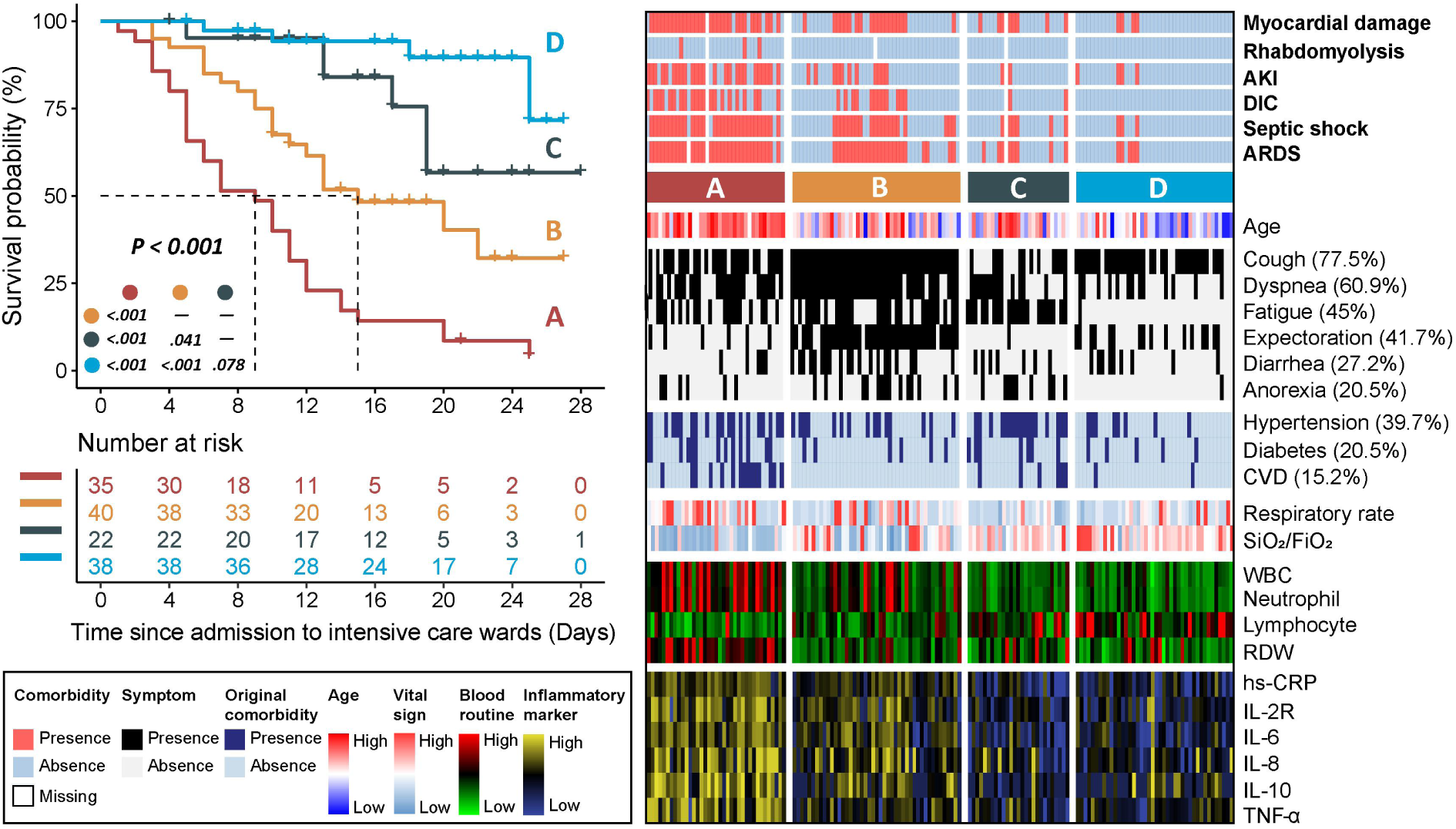
Clinical landscape of four prognostic types of COVID-19 in intensive care patients. Kaplan-Meier survival curves (left panel) showing differential survival rates; comprehensive heatmap (right panel) delineating clinical landscape of different types of COVID-19, with legend positioning in the left bottom panel. Survival was analyzed with log-rank test and pair-wise comparison was adjusted by Benjamini-Hochberg method. Labels of “high” and “low” were based on data interval instead of clinical reference values. AKI: acute kidney injury; DIC: disseminated intravascular coagulation; ARDS: acute respiratory distress syndrome; CVD: cardiovascular disease; SpO_2_: peripheral oxygen saturation; FiO_2_: fraction of inspired oxygen; WBC: white blood cell; RDW: red cell distribution width.

**Table.** Presenting characteristics of four types of COVID-19 in intensive care patients (n = 151)

|  | **A (n = 37)** | **B (n = 45)** | **C (n = 27)** | **D (n = 42)** |
| --- | --- | --- | --- | --- |
| Age, years | 77 (70-81) | 62 (52-70) | 65 (51-74) | 53 (43-58) |
| Signs and symptoms |  |  |  |  |
| Cough | 26 (70) | 45 (100) | 12 (44) | 34 (81) |
| Dyspnea | 24 (65) | 41 (91) | 14 (52) | 13 (31) |
| Fatigue | 19 (51) | 27 (60) | 21 (78) | 1 (2) |
| Expectoration | 6 (16) | 40 (89) | 1 (4) | 16 (38) |
| Diarrhea | 5 (14) | 20 (44) | 7 (26) | 9 (21) |
| Anorexia | 6 (16) | 15 (33) | 9 (33) | 1 (2) |
| Original comorbidities |  |  |  |  |
| Hypertension | 20 (54) | 15 (33) | 17 (63) | 8 (19) |
| Diabetes | 13 (35) | 6 (13) | 7 (26) | 5 (12) |
| CVD | 15 (41) | 0 | 6 (22) | 2 (5) |
| Vital signs |  |  |  |  |
| Respiratory rate, rpm | 25 (20-32) | 22 (20-26) | 20 (20-23) | 21 (20-25) |
| SpO<sub>2</sub>/FiO<sub>2</sub> | 99 (90-158) | 222 (100-294) | 297 (237-336) | 298 (248-345) |
| Laboratory findings |  |  |  |  |
| *Routine blood test* |  |  |  |  |
| WBCs, ×10<sup>9</sup>/L | 11.1 (8.0-15.5) | 7.4 (5.3-10.2) | 5.6 (4.5-6.5) | 5.2 (3.5-7.2) |
| Absolute lymphocytes, ×10<sup>9</sup>/L | 0.5 (0.3-0.7) | 0.7 (0.5-1.0) | 0.8 (0.7-1.2) | 1.0 (0.7-1.4) |
| Absolute Neutrophils, ×10<sup>9</sup>/L | 9.5 (7.1-15.0) | 5.5 (4.3-9.1) | 4.2 (3.0-5.1) | 3.1 (2.2-5.0) |
| RDW-CV | 13.4 (12.8-14.1) | 12.2 (11.9-12.8) | 12.6 (11.8-13.0) | 12.2 (11.8-12.7) |
| *Inflammatory marker* |  |  |  |  |
| hs-CRP, mg/L | 126 (76-190) | 80 (42-109) | 19 (5-49) | 28 (10-70) |
| IL-2R, U/ml | 1341 (940-1809) | 1038 (678-1185) | 701 (430-813) | 685 (439-928) |
| IL-6, pg/ml | 68 (37-137) | 43 (21-79) | 10 (2-20) | 14 (5-30) |
| IL-8, pg/ml | 42 (21-95) | 21 (13-40) | 12 (7-24) | 15 (10-21) |
| IL-10, pg/ml | 15 (9-24) | 7 (3-11) | 3 (3-6) | 3 (3-9) |
| TNF-α, pg/ml | 15 (10-23) | 9 (8-11) | 7 (5-9) | 8 (7-10) |

Continuous variables were described as median (IQR) while categorical variables were expressed as frequencies (%). All records were measured at admission to intensive care wards.
Multiple group comparison was done with Kruskal–Wallis test; proportions for categorical variables were compared using Fisher’s exact test. All calculated *P* values were less than 0.001 except for respiratory rate (*P*=0.004).
Abbreviations: CVD: cardiovascular disease; rpm: breaths per minute; SpO<sub>2</sub>: peripheral oxygen saturation; FiO<sub>2</sub>: fraction of inspired oxygen; WBC: white blood cell; RDW: red cell distribution width.

*Type A:* Extremely poor prognosis and elderly enriched; Dry cough, dyspnea and fatigue were common symptoms; hypertension, diabetes and CVD were common preexisting medical conditions. Patients presented severe respiratory failure, dramatically elevated counts of WBC and neutrophil, and lymphocyte depletion. Remarkable “cytokine storm” occurred, accompanied with later development of ARDS and multiple organ failure.

*Type B:* Poor prognosis and elderly enriched; dyspnea and cough with expectoration were common symptoms, accompanied with diarrhea and anorexia. Unfavorable respiratory condition and decreased lymphocyte count could be observed. Patients presented imminent “cytokine storm” and high risk of developing ARDS and multiple organ failure later after treatment.

*Type C:* Intermediate prognosis; symptoms of dry cough and fatigue, and original comorbidity of hypertension were common. Respiratory condition was normal and most laboratory tests were within normal or moderately elevated.

*Type D:* Favorable prognosis and middle age enriched; primary symptom was cough with expectoration. Patients had stable breathing and most laboratory tests were in normal range or slightly elevated.

## Discussion

This report, to our knowledge, is the first attempt of dealing with classification of COVID-19 in intensive care patients. The four types are prognosis-related and present a stepwise distribution in age, respiratory condition and inflammatory markers, suggesting the prognostic efficacy of these indicators. The specificity of symptoms does not appear to be strong, but gastrointestinal response (*e*.*g*., diarrhea) needs vigilance (4). Unexpectedly, hypertension is more evenly distributed, which contradicts previous study indicating hypertensive with COVID-19 was more likely to be in a high risk of mortality (5). Investigations in larger cohorts are required to provide more evidence.

The study is limited by ignoring the effect of treatment, but we believe that such classification of COVID-19 in intensive care patients could help in early warning of the disease and would provide effective triage for the treatment and management of individual patients, allowing for a more rational allocation of scarce medical resources in a resource constrained environment.

## Data Availability

Dr J. Wang had full access to all of the data in the study. After publication, the data will be made available to others on reasonable requests after approval from the author (J.W) and Wuhan Tongji Hospital (Y.L).

## Acknowledgments

We would like to thank all the hospital staff members for their efforts in collecting the information that was used in this study, and all the patients who consented to donate their data for analysis and the medical staff members who are on the front line of caring for patients.

## References

1. Guan W-j, Ni Z-y, Hu Y, Liang W-h, Ou C-q, He J-x, et al. Clinical Characteristics of Coronavirus Disease 2019 in China. N Engl J Med. 2020. doi: 10.1056/NEJMoa2002032.

2. Mo Q, Shen R, Guo C, Vannucci M, Chan KS, Hilsenbeck SG. A fully Bayesian latent variable model for integrative clustering analysis of multi-type omics data. Biostatistics. 2018;19(1):71–86. doi: 10.1093/biostatistics/kxx017.

3. Yang X, Yu Y, Xu J, Shu H, Liu H, Wu Y, et al. Clinical course and outcomes of critically ill patients with SARS-CoV-2 pneumonia in Wuhan, China: a single-centered, retrospective, observational study. Lancet Respir Med. 2020. doi: 10.1016/S2213-2600(20)30079-5.

4. Liang W, Feng Z, Rao S, Xiao C, Xue X, Lin Z, et al. Diarrhoea may be underestimated: a missing link in 2019 novel coronavirus. Gut. 2020. doi: 10.1136/gutjnl-2020-320832

5. Zhou F, Yu T, Du R, Fan G, Liu Y, Liu Z, et al. Clinical course and risk factors for mortality of adult inpatients with COVID-19 in Wuhan, China: a retrospective cohort study. Lancet. 2020. doi: 10.1016/S0140-6736(20)30566-3

